# Outcome of complex surgical resection and reconstruction for rare thoracic cancers: the clinical value of a predictive score

**DOI:** 10.1101/2022.05.11.22274955

**Authors:** Ugo Pastorino, Giovanni Leuzzi, Federica Sabia, Paolo Girotti, Leonardo Duranti, Stefano Radaelli, Marco Fiore, Silvia Stacchiotti, Giannatempo Patrizia, Roberto Salvioni, Alessandro Gronchi

## Abstract

**Background:** Complex surgical resection and reconstruction for rare thoracic cancers (RTCs) represent a major challenge, given their very low frequency, extreme variability of presentation, multi-modality treatment options and inadequate outcome prediction. We reported the experience of a tertiary referral centre on a consecutive series of RTC patients, to predict outcome by disease and complexity of surgical procedures.

**Methods:** From Jan 2003 to Dec 2018, 1122 surgical procedures were performed with curative intent on 952 RTC patients. Study endpoints were: post-operative hospital stay (Pod), 30-day and 90-day mortality, 5-year and 10-year survival (OS). The follow-up was closed at June 2020.

**Results:** Median Pod was 8 days, with a 2% 30-day and 3.9% 90-day mortality. Overall survival (OS) was 85.7% at 1 year, 61.7% at 5 years and 50.7% at 10 years. Ten-year OS was 64.8% in low, 58.8% in intermediate, and 42.4% in high complexity score (Log-rank tests p<0.0001); 64.4% in patients with 1 or 2 reconstructions and 32.8% in patients with 3 or more reconstructions; 44.5% with vascular and 48% with chest wall reconstruction; 71.8% in germ cell tumors and 0% in mesothelioma.

**Conclusion:** Complex surgical resection and reconstruction was associated with acceptable 90-day mortality and good 10-year survival in all RTCs but mesothelioma.

A predictive score based on surgical complexity and cancer type can help the clinical decision making.

## INTRODUCTION

Rare cancers encompass a very heterogeneous group of diseases with an incidence of fewer than 6 cases per 100,000 individuals per year [1] and accounting about 25% of all malignancies worldwide [2]. Specifically, rare thoracic cancers (RTCs) represent 8% of all tumours of the chest cavity [3] with a crude incidence in Europe and in Italy of 6.8 and 5.4 per 100 000 people per year, respectively [2,3]. According to the results of RARECAREnet and SEER databases, in European Union (EU) and USA the 5-year survival of all rare cancers is lower (48.5 and 55%, respectively) compared with all common cancers (63.4 and 74%, respectively) [2,3]. The reasons for this worse survival may be related to the lack of effective and standardized treatments as well as delays in diagnosis [4]. On the other hand, RTCs represent an exception in terms of survival, confirming a better outcome in EU and Italian population (13.4% and 17%, respectively) compared with their more common counterparts with a 5-year survival of 10.1% [2,3].

In the setting of RTCs, the SEER database (from 2009 to 2013) reports an incidence of 0.22-0.25 cases per 100,000 per year for thymomas, 0.04-0.09 for mediastinal germ-cell tumors, 0.01-5.30 for oesophageal carcinomas, 0.03-0.80 for thoracic sarcomas and 0.30-1.49 for malignant pleural mesothelioma (MPM) [1]. Such a low incidence causes a limited source of data from the literature, with small case series [5-7], no prospective studies [8-10] and very rare registry studies [11-15] reported. Given their very low frequency, extreme variability of clinical presentation, different treatment options and limited individual expertise, RTCs represent a challenge for most surgeons that could face with troubles in surgical decision making and choice of multimodality strategy. The available literature indicates the need to centralize diagnostic work-up and treatment in reference centres with high volume, specific expertise, and multidisciplinary approach.

In this scenario, we reported our experience of a tertiary referral centre on an unselected series of RTC patients to provide more solid evidence on surgical management, perioperative and long-term outcome with a new predictive model, based on the complexity of surgical procedures.

## METHODS

### Study design and participants

From January 2003 to December 2018, 1265 consecutive patients admitted to the IRCCS Istituto Nazionale dei Tumori of Milan with a diagnosis of RTC were retrospectively evaluated, and 952 undergoing a curative resection were included in this analysis, 795 for primary tumor and 157 for metastatic disease. All available information on tumor type, description of surgery (date, type, complexity, number and type of reconstructions), post-operative stay and follow-up were retrieved. Among metastatic patients, only thymomas or germ-cell tumors with pleural or mediastinal metastases were included.

Complexity of surgical procedure was classified in Low, Intermediate and High. (as reported in the Supplementary Material). Patients with unresectable tumor, macroscopic (R2) residual resection or undergoing biopsy alone were excluded from the analysis. Demographic information was collected at the hospitalization and the vital status was obtained through the Istituto Nazionale di Statistica (ISTAT, SIATEL 2.0 platform), which provides the exact date of death within 3 months of occurrence. The follow-up was closed at June 2020.

The study was approved by the appropriate institutional review committee and meet the guidelines of their responsible governmental agency.

### Statistical analysis

The study endpoints were: a) post-operative hospital stay; b) post-operative 30-day and 90-day mortality; c) post-operative 1-year, 5-year and 10-year survival (OS). Follow-up time was calculated from the date of surgery to the date of death from any cause or last follow-up. Proportions were compared by Chi-square test or Fisher’s exact test, as appropriate. Survival functions were estimated using Kaplan–Meier Curves and comparisons were tested by Log-rank test. Multivariate Cox regression models were performed to estimate hazard ratios (HRs) and 95% Confidence Interval (CI) of complexity score and number of reconstructions, adjusted for age, sex and type of tumor. Long-term survival curves were truncated at 10 years. All analyses were performed using the Statistical Analysis System Software (Release SAS:9.04; SAS Institute, Cary, North Carolina, USA).

## RESULTS

### Patient characteristics and surgical information

A total of 1508 surgical procedures for rare thoracic tumors were performed from January 2003 to December 2018, 1122 (74.4%) procedures on 952 patients were curative resections and 386 (25.6%) procedures on 313 patients were biopsies (Figure 1). All patients treated with curative intent were included in this analysis: 795 (83.5%) resections for primary tumor and 157 (16.5%) for metastatic disease.

**Figure 1.**
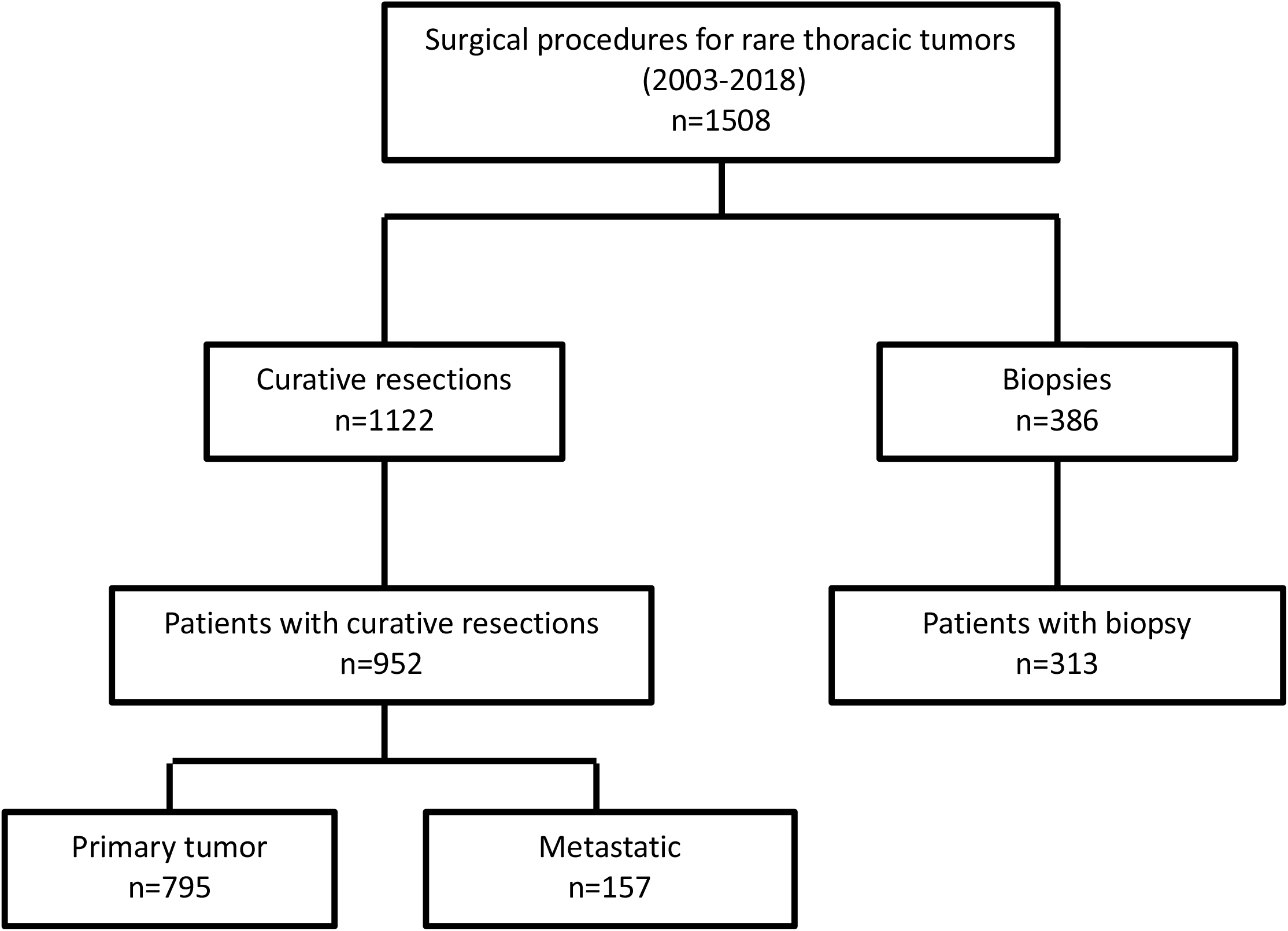
Consort diagram

Patient’s characteristics are summarized in Table 1: median age was 55 years (IQR 39-65) and 33.8% were female. Patients with germ cell tumor were younger than other types, with median age of 32 (IQR 26-39) and most of them were males (195/205, 95%). Conversely, most of the older patients (>=65 years) had esophageal carcinoma (29%), thymoma (25.3%) or sarcoma (23.4%). Females were mostly affected by thymoma (31.4%) or sarcoma (32%), and 64.6% had a high complexity score. Complexity score and number of reconstructions were higher in older patients. Details on the numbers of reconstructions made according to tumor type were reported in Table S1. Esophageal carcinoma accounted for 39.6% of patients with 3 ore more reconstructions. The median age of patients who underwent vascular reconstructions was 45.5, and of chest wall reconstruction 55.5.

**Table 1:**
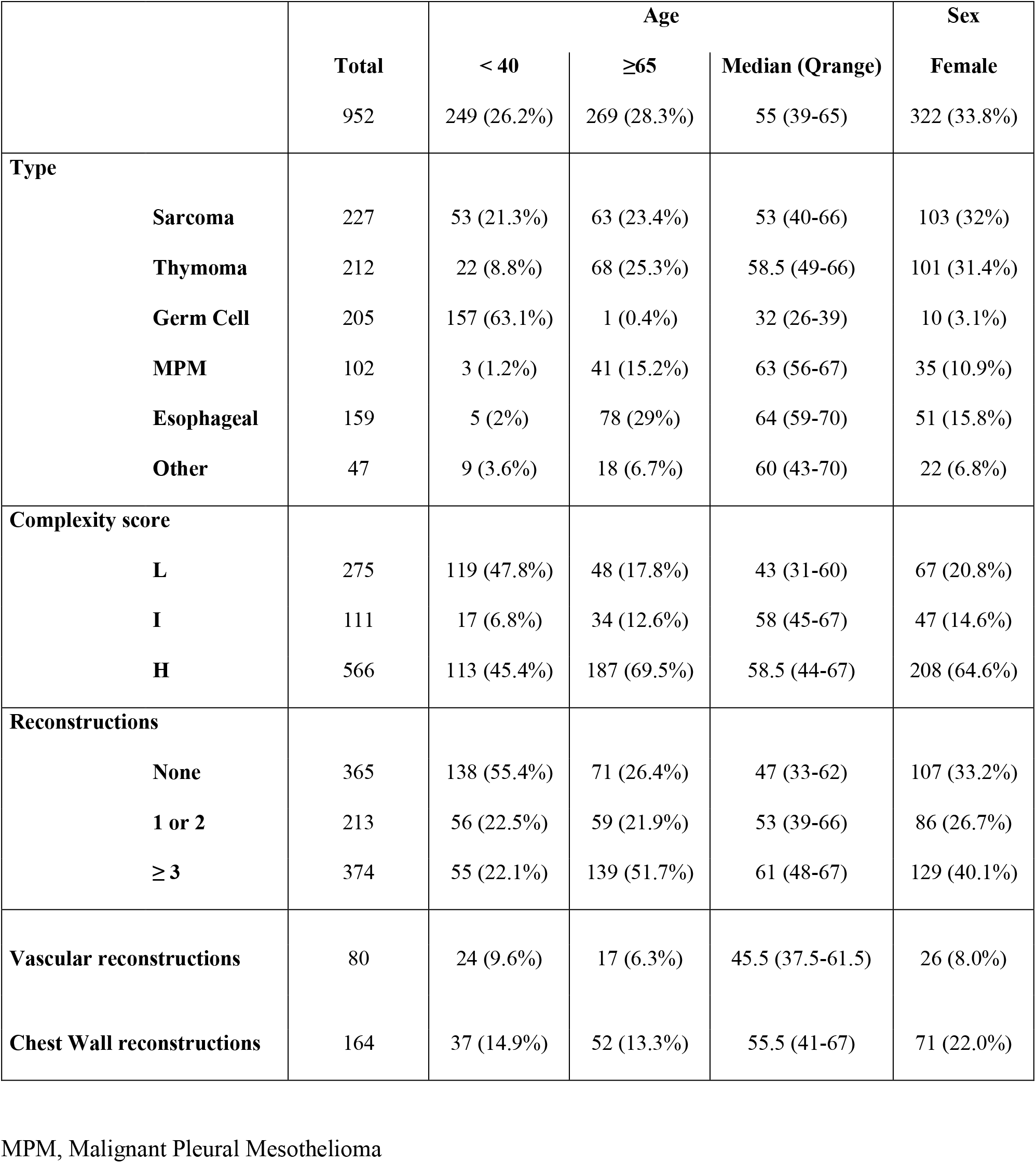
Patients’ characteristics according to tumor and surgical information.

### Post-operative mortality

In all 1122 resections, median post-operative hospital stay (Pod) was 8 days, with a 2% 30-day mortality and 3.9% 90-day mortality (Table 2).

**Table 2:** Postoperative hospital stay and mortality in 1122 resections (of 952 patients)

|  |  | N resections | Pod<br>(median) | 30d MO | P-value | 90d MO | P-value |
| --- | --- | --- | --- | --- | --- | --- | --- |
| **Total** |  | 1122 | 8 | 2.0% |  | 3.9% |  |
| **Type** | **Sarcoma** | 259 | 8 | 1.5% | 0.0357<sup>a</sup> | 2.7% | 0.1636<sup>a</sup> |
|  | **Thymoma** | 237 | 6 | 1.7% |  | 3.0% |  |
|  | **Germ Cell** | 272 | 6 | 0.4% |  | 2.9% |  |
|  | **MPM** | 118 | 11 | 2.5% |  | 5.9% |  |
|  | **Esophageal** | 182 | 14 | 4.4% |  | 6.0% |  |
|  | **Other** | 54 | 7 | 3.7% |  | 7.4% |  |
| **Complexity score** | **L** | 389 | 5 | 0.5% | 0.0059<sup>a</sup> | 2.3% | 0.0347<sup>a</sup> |
|  | **I** | 134 | 6.5 | 0.8% |  | 2.2% |  |
|  | **H** | 599 | 11 | 3.2% |  | 5.3% |  |
| **Reconstructions** | **None** | 498 | 6 | 1.0% | 0.0081<sup>a</sup> | 2.6% | 0.0100<sup>b</sup> |
|  | **1 OR 2** | 227 | 7 | 0.9% |  | 2.6% |  |
|  | **≥ 3** | 397 | 13 | 3.8% |  | 6.3% |  |
| **Vascular reconstructions** |  | 86 | 12 | 5.8% |  | 10.4% |  |
| **Chest Wall reconstructions** |  | 172 | 9 | 1.2% |  | 3.5% |  |

MPM, Malignant Pleural Mesothelioma; Pod, post-operative day; MO, mortality.
<sup>a</sup> Fisher exact test
<sup>b</sup> Chi-Squared test

The median number of Pod was the highest in esophageal carcinomas (14 days) and the lowest in thymoma and germ cell tumors (6 days). Pod increased with the increase of complexity and of the number of reconstructions. Statistically significant difference in 30-day mortality was found in tumor types (p=0.0357): 0.4% in germ cell tumor, 1.5% in sarcoma, 1.7% in thymoma, 2.5% in MPM, 4.4% in esophageal carcinoma, and 3.7% in other types. A non-statistically significant difference was found in 90-days mortality (p=0.1636): 2.7% in sarcoma, 2.9% in germ cell tumor, 3.0% in thymoma, 5.9% in MPM, 6.0% in esophageal carcinoma, and 7.4% in other types. Both 30- and 90-days mortalities were statistically different in strata of complexity score (p=0.0059 and p=0.0347 respectively): resections with a high complexity score had a higher 30-days mortality (3.2%) as well as a higher 90-days mortality (5.3%) compared to low or intermediate score. Similarly, statistically significant differences were found stratifying by the number of reconstructions (30-days p=0.0081, 90-days p=0.0100): patients with ≥3 reconstructions had a higher 30- (3.8%) and 90-days mortality (6.3%) compared to none (1.0% and 2.6% respectively) and ≤2 reconstructions (0.9% and 2.6% respectively). The results of multivariate Cox models stratified by complexity score and number of reconstructions (Table S2) showed a significantly higher risk of 30-day post-operative mortality in resections with a high complexity score (HR 6.54, p=0.0460) compared to low complexity score in Model A with the adjustment for age and sex. After the further adjustment for tumor type, a high complexity score maintained a borderline significant higher risk (HR=6.33, p=0.0578, Model B).

### Long-term survival

Overall survival (OS) was 85.7% at 1 year, 61.7% at 5 years and 50.7% at 10 year (Table 3). OS was significantly different among tumor types, with Log-rank test p<0.001 at 1, 5 and 10 years (Table 3). 1- and 5-year OS were better in thymoma (96.2% and 84.8%) and in germ cell tumor (90.7% and 75.7%), while MPM had the worst outcomes (70.2% and 21.7% respectively). 10-year OS was better in germ cell tumors (71.8%), followed by thymomas (64.6%), sarcomas (51.3%), other types (49.5%), esophageal carcinomas (31.2%), and lastly MPM (0%) (Table 3 and Figure 2 panel A). More complex surgery was associated with a worst OS, at 1, 5 and 10 years. 10-year OS was statistically different in levels of complexity score (Log-rank tests p<0.0001): 64.8% in low, 58.8% in intermediate, and 42.4% in high complexity score (Table 3 and Figure 2 panel B). Similarly, OS was worst in patients undergoing the highest number of reconstructions, at 1, 5 and 10-years (Log-rank tests p<0.0001). 10-year OS was 32.8% in patients with 3 or more reconstructions, 64.4% in patients with 1 or 2 reconstructions and 62.1% in patients with no reconstructions (Table 3 and Figure S1). 10-year OS was 44.5% in patients undergoing vascular reconstructions and 48% in chest wall reconstructions (Table 3).

**Table 3:** Cumulative survival at 1 year, 5 years and 10 years of 952 patients.

|  |  | N<br>patients | 1-year<br>cumulative<br>survival | Log-Rank<br>P-value | 5-year<br>Cumulative<br>survival | Log-Rank<br>P-value | 10-year<br>cumulative<br>survival | Log-Rank<br>P-value |
| --- | --- | --- | --- | --- | --- | --- | --- | --- |
| **Total** |  | 952 | 85.7% |  | 61.7% |  | 50.7% |  |
| **Type** | **Sarcoma** | 227 | 85.8% | <0.0001 | 63.1% | <0.0001 | 51.3% | <0.0001 |
|  | **Thymoma** | 212 | 96.2% |  | 84.8% |  | 64.6% |  |
|  | **Germ Cell** | 205 | 90.7% |  | 75.7% |  | 71.8% |  |
|  | **MPM** | 102 | 70.6% |  | 21.7% |  | 0% |  |
|  | **Esophageal** | 159 | 76.0% |  | 37.3% |  | 31.2% |  |
|  | **Other** | 47 | 80.9% |  | 62.3% |  | 49.5% |  |
| **Complexity score** |  |  |  |  |  |  |  |  |
|  | **L** | 275 | 93.8% | <0.0001 | 75.1% | <0.0001 | 64.8% | <0.0001 |
|  | **I** | 111 | 92.8% |  | 73.2% |  | 58.8% |  |
|  | **H** | 566 | 80.3% |  | 53.0% |  | 42.4% |  |
| **Reconstructions** |  |  |  |  |  |  |  |  |
|  | **None** | 365 | 92.9% | <0.0001 | 72.5% | <0.0001 | 62.1% | <0.0001 |
|  | **1 OR 2** | 213 | 90.1% |  | 75.4% |  | 64.4% |  |
|  | **≥ 3** | 374 | 76.1% |  | 43.7% |  | 32.8% |  |
| **Vascular reconstructions** |  | 80 | 80.0% |  | 63.6% |  | 44.5% |  |
| **Chest Wall reconstructions** |  | 164 | 84.1% |  | 57.0% |  | 48.0% |  |

MPM, Malignant Pleural Mesothelioma

**Figure 2 Panel A:**
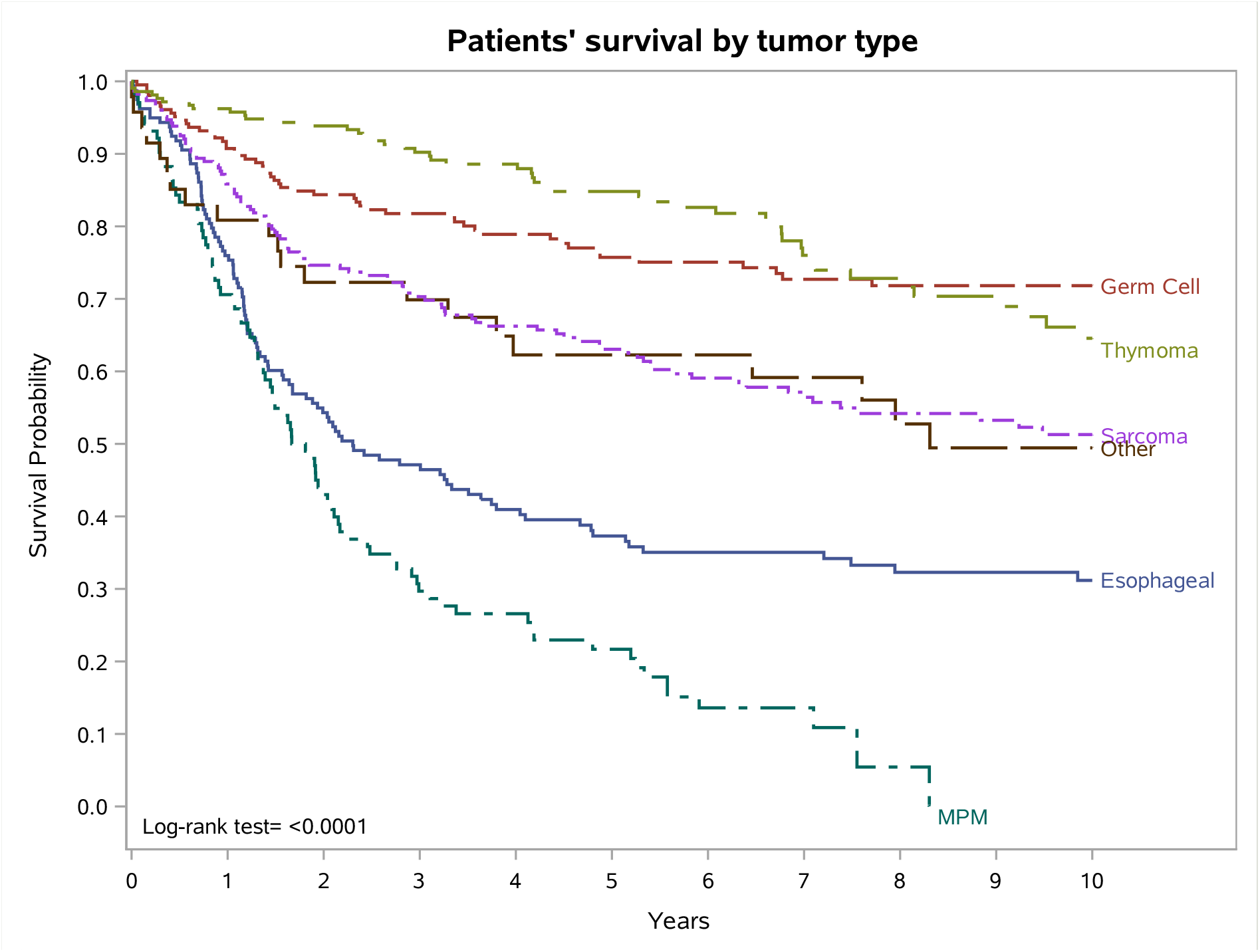
Patients’ survival by tumor type.

**Figure 2 Panel b.**
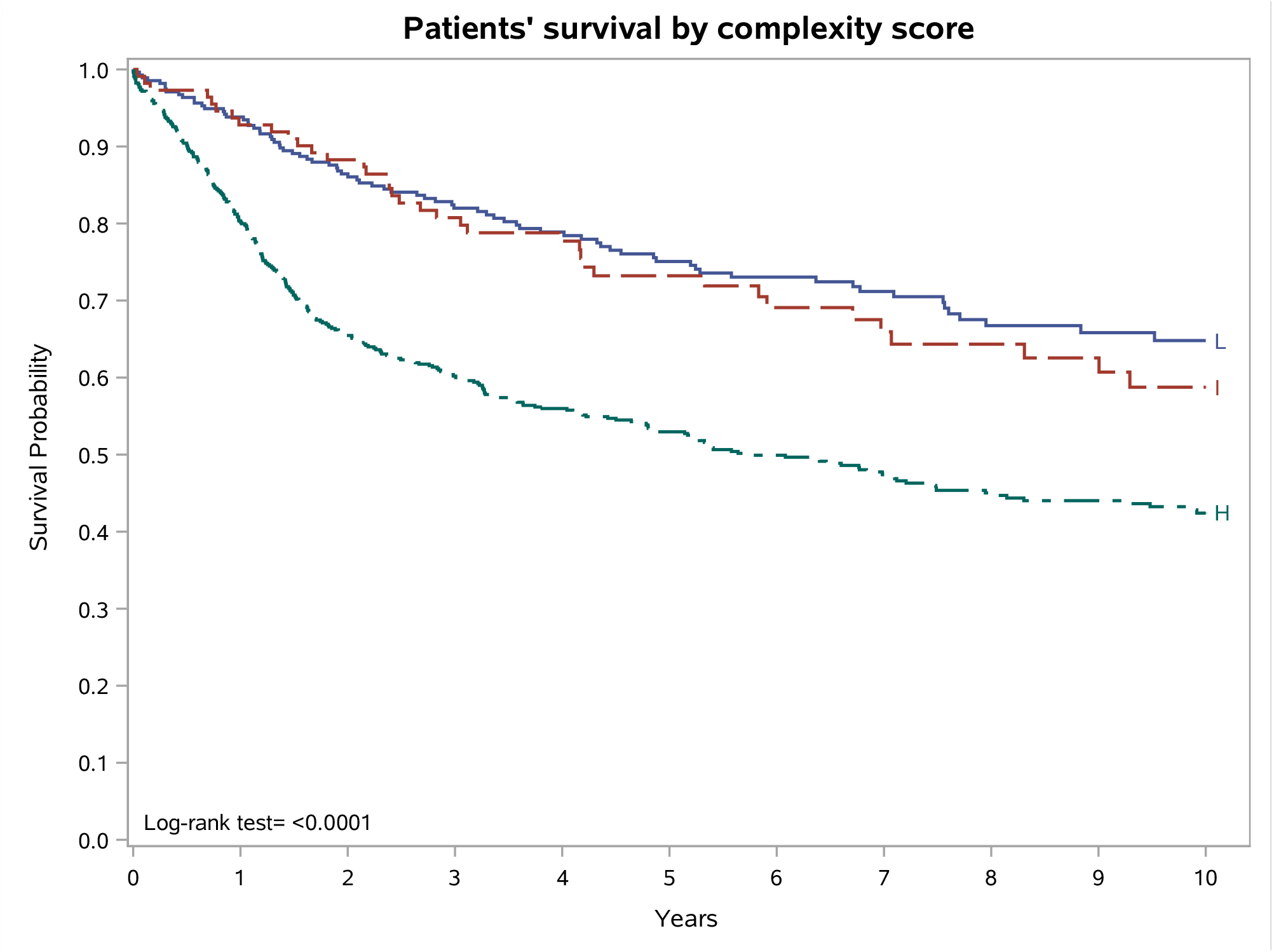
Patients’ survival by complexity score

Kaplan-Meier curves restricted to patients with 3 or more reconstructions showed 10-year OS in tumor types similar to that described above for all subjects, except for MPM and other tumor types which showed a much lower survival than the overall analysis (Figure 3). 10-year survival curves of patients with any reconstructions stratified by tumor type are reported in Figure S2. For germ cell tumors and for thymomas, Table S3 shows the comparisons of 5-year and 10-year survival in primary and in metastatic disease, overall and stratifying by complexity score. For germ cell tumor 5- and 10-year OS were respectively 75% and 73% in primary, 76% and 71% in metastatic disease. For thymoma 5- and 10-year OS were respectively 87% and 65% in primary, 63% and 52% in metastatic disease. The results of age and sex adjusted multivariate Cox models (Table S4) showed a nonsignificant impact of complexity score on 5-year survival (HR 1.53, p=0.0604) and a statistically significant higher risk in patients with 3 or more reconstructions (HR 1.66, p=0.0126, Model A). After the further adjustment for tumor type, the high complexity score had a significant higher risk (HR=2.66, p<0.0001, Model B) compared to low score. A similar risk profile was observed for 10-year survival (HR 1.36, p=0.1345, Model A, and HR=2.21, p=0.0001, Model B). Patients with 3 or more reconstructions had a significant lower 10-year survival (HR 1.75, p=0.0029, Model A), but the difference lost significance after adjustment for tumor type (HR 1.10, p=0.6233, Model B).

**Figure 3.**
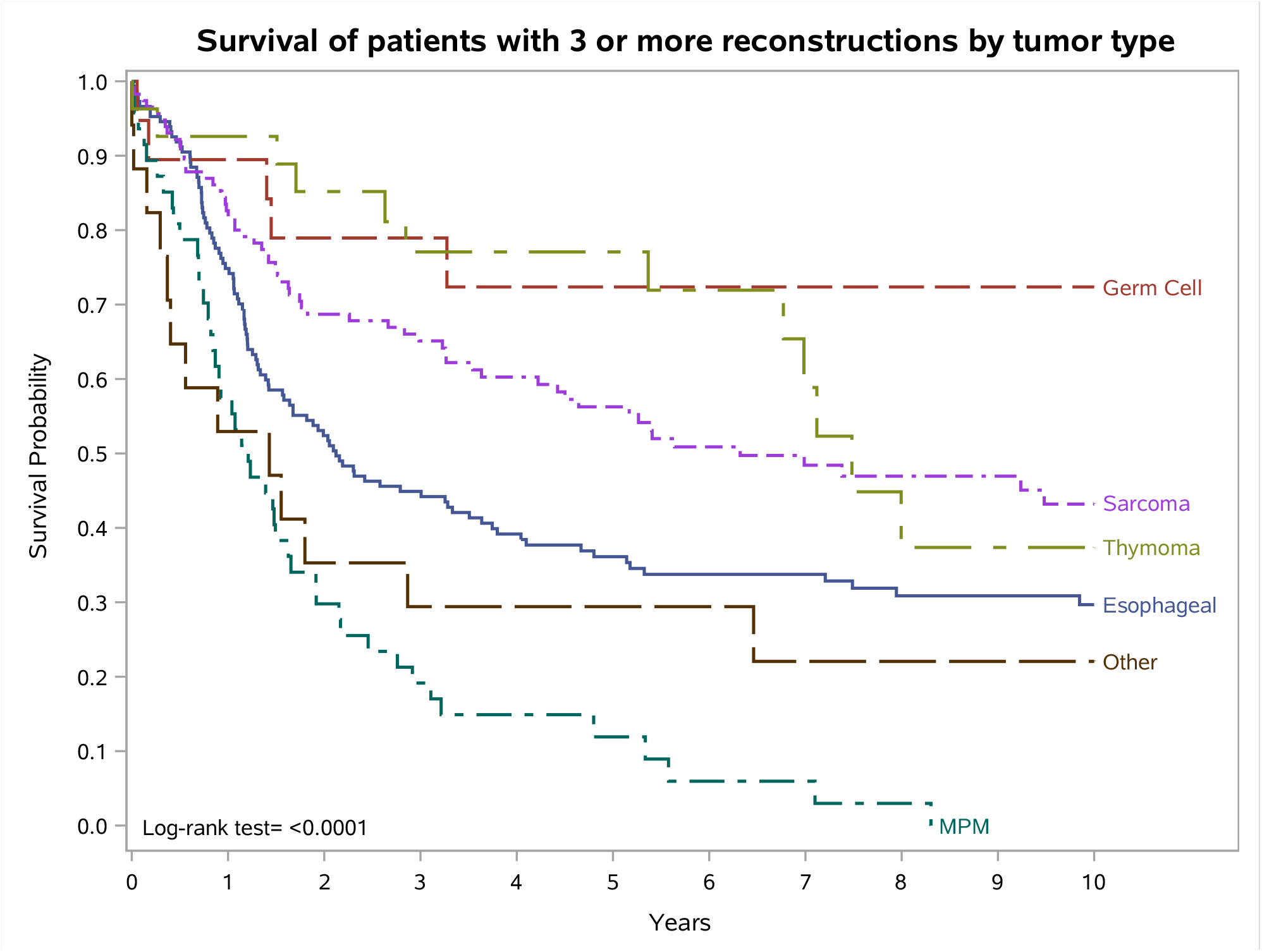
Survival of patients with 3 or more reconstructions by tumor type

## DISCUSSION

Tumor-Node-Metastasis (TNM) staging system was the first oncological predictive model proposed in 1953 by Pierre Denoix as a simple universal language to predict solid “common” tumour prognosis [16]. After many years, TNM system remains the gold standard for all clinicians, notwithstanding well-known limitations. In fact, as reported by Balachandran VP, TNM “is rooted in the Halstedian principle of temporal determinism that solid tumours spread sequentially from the primary site to lymphatics, then to distant organs, categorizing patients by anatomic spread of disease and survival” [17]. This “extreme” simplification and categorization into limited number of stages may lead to high heterogeneity as well as to difficulties in predicting individual patient’s prognosis [17]. To overcome these biases, a lot of tumour-specific nomograms have been proposed, taking into account either preoperative or postoperative cancer/patients’ features. Furthermore, given the worldwide availability of large patients database, the definition and construction of new prognostic models may appear relatively “simple” in the setting of common solid tumours. Unfortunately, this is not applicable to the subset of rare cancers.

Contrary to the small case series reported in literature, we analysed a large monocentric database of unselected RTC patients and, to our knowledge, reported for the first time a predictive analysis that estimated the long-term outcome. In the literature, prognostic models are rarely reported in this setting and, specifically, only for single rare diseases such as for primary mediastinal germ-cell tumours [18], thymoma [15], oesophageal carcinoma [19] and MPM [14, 20], while no predictive models (except for their retroperitoneal counterparts [21]) have been proposed for thoracic sarcomas. In addition, these scores mainly consider clinical or pathological features, leaving out the “real” surgical impact on natural history of these neoplasms. To overcome the high heterogeneity of RTC diseases, we grouped patients according to the extension (and complexity) of surgical procedures. As shown in Figure 2 panel B, patients undergoing single (not-combined) resections (i.e., lobectomy, pleurectomy) had the best 10-year survival (64.8%), but even patients with extensive resections (H) showed a favourable outcome (42.4%). The prognostic value of the extension (and complexity) of surgical procedures in post-operative mortality and long-term survival was also confirmed by multivariate analysis (Table S2 and S4), although significance at level 0.05 was not always reached, due to the relatively small numbers in each subgroup.

In the literature, the prognostic impact of surgical extension has been rarely addressed, and only in terms of early outcome [22]. In the context of rare cancers, where specific staging systems are not comparable, this surgical complexity score may represent a simpler tool to categorize patients by anatomical extension, analogously to TNM system. This evidence is further supported by survival analyses according to the extent of reconstructions (Figure 3), where the degree of surgical invasiveness predicts the outcome of RTCs. In clinical practice, these results provide a valuable support to multidisciplinary preoperative evaluation (radiological and functional) of these rare patients, and guidance in the decision-making process. On the other hand, they confirm the need of RTC referral to highly qualified centres, to select who may benefit from extensive surgery, predict the outcome (by using the complexity score) and apply the appropriate intraoperative and postoperative management. As a matter of fact, when performed in high-volume centres, complex operations can be carried out with satisfactory outcomes.

The role of vascular invasiveness and related surgery (resection and vascular reconstruction) in RTCs has also a matter of debate over the past years. Especially for advanced thymic and germ-cell neoplasms, infiltration of mediastinal great vessels (innominate veins and superior vena cava, SVC) is common. In this context, vascular infiltration increases the technical complexity to achieve complete resection at the vascular site and requires an effective reconstructive strategy. In primary lung cancer (NSCLC), many surgeons can mention some bad experiences when facing involvement of innominate veins or SVC, and poor survival even in the case of successful vascular reconstruction, with a 5-year OS ranging from 15 to 40% [23-25]. On the contrary, few authors have reported interesting outcomes in vascular resections for thymic neoplasms. Comacchio et al. reported a favourable outcome in 144 vascular resections for thymomas (5- and 10-years OS rates of 75% and 56%, respectively), with a worse DFS in case of pathological vascular infiltration (49% and 41%, respectively) [26]. Similarly, a retrospective analysis performed by Yu Z et al [27] reported a 5- and 10-year OS of 93.94%, and 60.81% in a series of 45 patients undergoing vascular resection/reconstruction. Compared to NSCLC patients, RTCs outcome reflects the specific tumour biology as well as global patients’ features: younger age, less risk factors (smoking) and/or comorbidities (emphysema, coronary disease). Noteworthy, given the small numbers, it is hard to find a comparison of outcome in different RTCs undergoing vascular reconstruction.

Our results confirmed the acceptable long-term survival (5- and 10-years OS rates of 63.6% and 44.5%, respectively), and gave us the opportunity to perform a “homogeneous” evaluation of the role of extensive surgery in the various RTC types, with or without vascular invasion. We are aware that further prospective investigations are needed to better understand this issue.

The results of our study also confirmed those reported by RARECAREnet and AIRTUM database for RTCs [2,3], showing the best prognosis for resected thymomas, primary mediastinal germ-cell tumours and thoracic sarcoma, with a 5-yr survival of 84.8%, 75.7% and 63.1%, respectively.

Furthermore, our study underlines the role of surgery as a part of multimodality therapy in germ-cell tumours, where the cure rate strongly depends on setting and first-line chemotherapy (*cisplatin*-based *combination chemotherapy*) as well as on complete resection of residual mass [28], due to the risk of persistent viable cells, teratoma and/or teratoma with malignant transformation [10, 29]. In mediastinal germ-cell tumours, surgery has proven to improve cure rates [11, 30] and predict survival by post-chemotherapy pathologic assessment [31]. Our results demonstrate that surgery for primary mediastinal germ-cell tumours is associated with a very favourable outcome even in patients with high complexity score (10-yr OS of 72%, Table S3).

Conversely, the role of surgery in MPM is still strongly debated. MPM is associated to very dismal outcome with a 5-yr survival ranging from 4.6% in Europe to 7% in Italy [2,3]. To date, no consensus has been yet reached regarding the gold-standard for surgical patients if considering that the MARS 1 trial [32] increased only our doubts regarding the usefulness of extra pleural-pneumonectomy (EPP) for MPM patients. These results are in line with that reported in a recent meta-analysis on over 3900 mesothelioma patients confirming that P/D is associated with enhanced outcomes regarding 30-day mortality, median overall survival, and complications [33]. Furthermore, the results of MesoVATS (the second-ever randomized controlled trial on surgical management of MPM) [34] revealed that palliative surgery (talc pleurodesis with biopsy) was preferable to partial pleurectomy, further underlying the hypothesis that “cytoreduction”, as well extensive thoracic demolitions, may be ineffective for MPM [34]. Such a dismal prospect is confirmed by our selected population of 102 MPMs resected with curative intent, whose survival is by far the worst among all RTC. New evidence on this interesting issue could be generated by the on-going MARS 2 trial [35].

In conclusion, the long-term survival reported in our unselected RTCs series, demonstrates that a careful preoperative evaluation combined with specific expertise in complex surgical resections and a multidisciplinary management provide the best chance cure, especially for primary mediastinal germ-cell tumours, thymomas and sarcomas. This is not applicable to MPM, in which the gold standard of therapy has not been defined yet. We hope that in the future it may be more feasible to share data on these rare diseases, to predict prognosis and define the best “tailored” treatment. A predictive score based on surgical complexity and cancer type can help the clinical decision making.

## Data Availability

All data produced in the present study are available upon reasonable request to the authors

