## Supplementary Material for "Outcome of complex surgical resection and reconstruction for rare thoracic cancers: the clinical value of a predictive score"

### Complexity score

All procedures were classified in three levels, according to their complexity:
low , intermediate and high.

| | |
|---|---|
| Low complexity score (**L**): | simple mediastinal resection<br>endothoracic lymphadenectomy<br>sublobar polmonary resection<br>lobectomy / segmentectomy<br>pleurectomy and/or decortication<br>partial diaphragmatic resection<br>partial rib resection. |
| Intermediate complexity score (**I**): | radical tymectomy<br>vascular resection<br>left pneumonectomy<br>sleeve lobectomy / segmentectomy<br>chest-wall resection < 3 ribs<br>partial sternectomy<br>diaphragmatic reconstruction<br>myo-cutaneous flap / omentoplasty |
| High complexity score (**H**): | extended tymectomy<br>esophagectomy<br>vascular reconstruction / prosthetic replacement<br>right pneumonectomy<br>extra-pleural / completion pneumonectomy<br>sub-total / total sternectomy<br>chest-wall resection ≥ 3 ribs with rigid prosthesis<br>vertebral resection<br>thoraco-pleuro-pneumonectomy |

**Table S1.** Number of reconstructions according to tumor type.

| | 0 reconstruction | 1 or 2 reconstructions | ≥ 3 reconstructions | TOTAL |
|---|---|---|---|---|
| **Sarcoma** | 62 (17.0%) | 49 (23.0%) | 116 (31%) | 227 |
| **Thymoma** | 75 (20.6%) | 110 (51.6%) | 27 (7.2%) | 212 |
| **Germ Cell** | 160 (43.8%) | 26 (12.2%) | 19 (5.1%) | 205 |
| **MPM** | 35 (9.6%) | 20 (9.4%) | 47 (12.6%) | 102 |
| **Esophageal** | 10 (2.7%) | 1 (0.5%) | 148 (39.6%) | 159 |
| **Other** | 23 (6.3%) | 7 (3.3%) | 17 (4.6%) | 47 |
| **Total** | 365 | 213 | 374 | 952 |

MPM, Malignant Pleural Mesothelioma

**Figure S1**. Patients' survival according to the number of reconstructions.

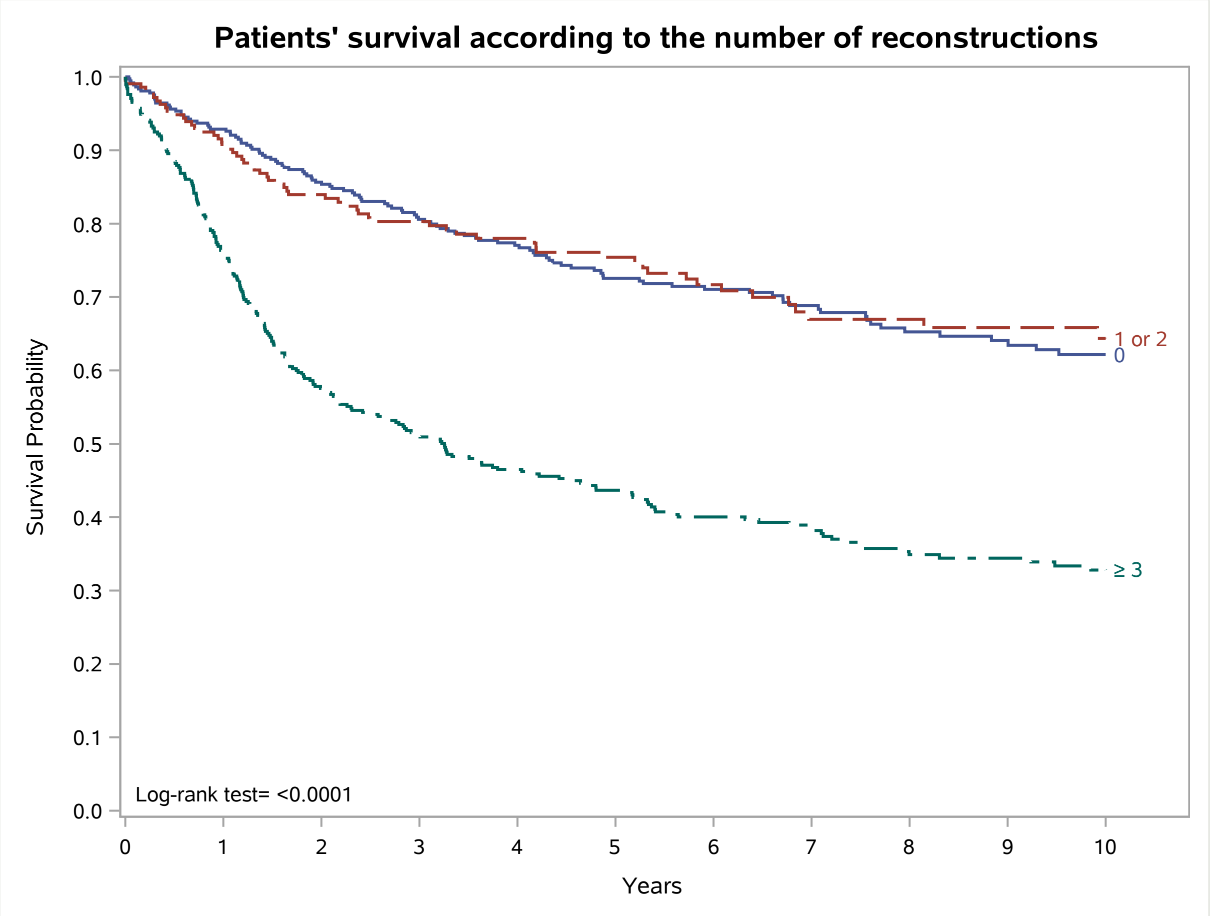

**Figure S2**. Survival of patients with any reconstructions according to tumor type.

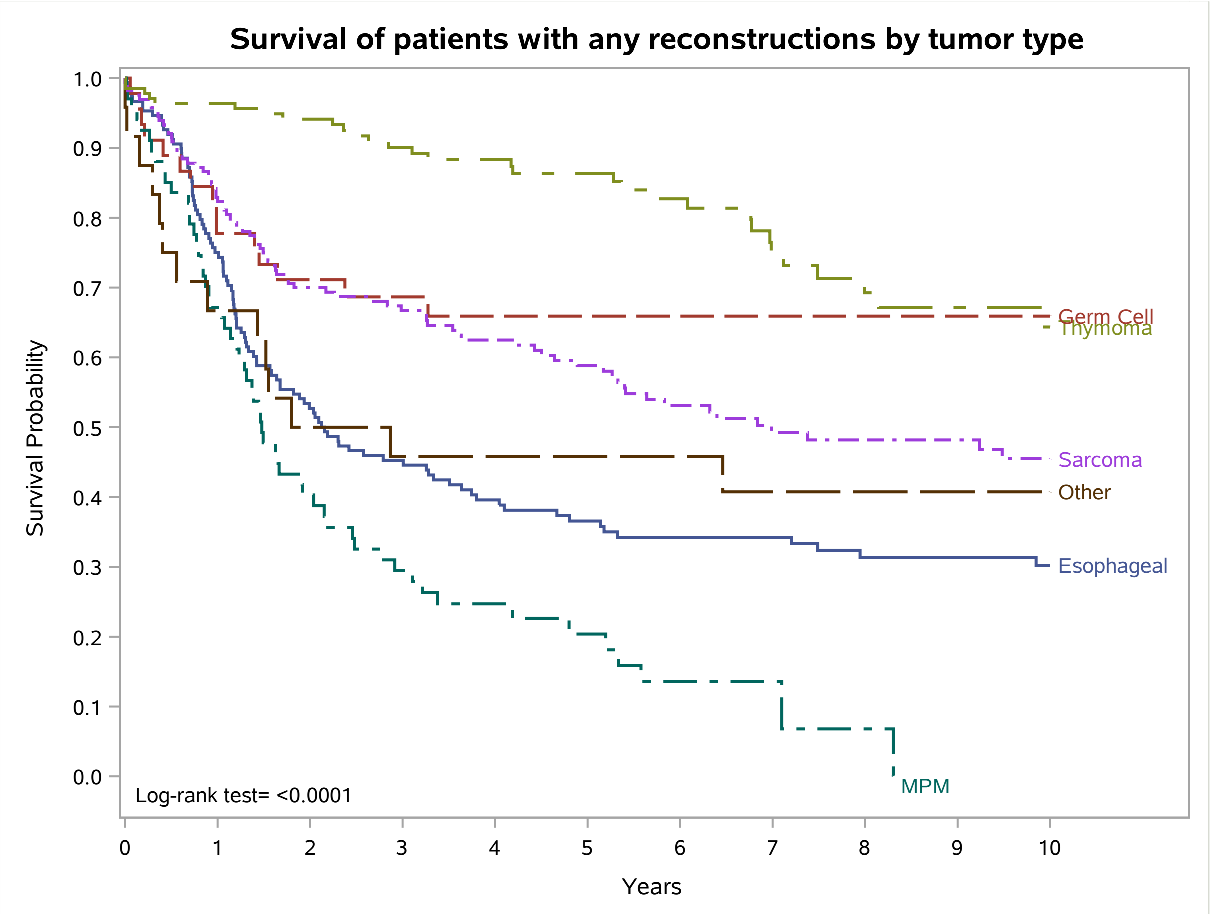

**Table S2**: Multivariate Cox regression models for 30 day and 90 day post-operative mortality: Model A adjusted for age and sex ; Model B adjusted for age, sex and type of tumor.

| | **30d post-operative mortality** *HR (95%CI)* *p-value* | | **90d post-operative mortality** *HR (95%CI)* *p-value* | |
|---|---|---|---|---|
| | **Model A** | **Model B** | **Model A** | **Model B** |
| **Complexity score** | | | | |
| **L** | Ref. | Ref. | Ref. | Ref. |
| **I** | 1.46 (0.13-16.68)<br>0.7613 | 1.31 (0.11-15.46)<br>0.8297 | 0.97 (0.25-3.86)<br>0.9689 | 1.18 (0.29-4.84)<br>0.8144 |
| **H** | **6.54 (1.03-41.39)**<br>**0.0460** | 6.33 (0.94-42.58)<br>0.0578 | 1.94 (0.58-6.50)<br>0.2815 | 2.59 (0.76-8.86)<br>0.1294 |
| **Reconstructions** | | | | |
| **None** | Ref. | Ref. | Ref. | Ref. |
| **1 OR 2** | 0.28 (0.05-1.69)<br>0.1634 | 0.27 (0.04-1.68)<br>0.1593 | 0.66 (0.19-2.22)<br>0.4963 | 0.73 (0.21-2.46)<br>0.6079 |
| **≥ 3** | 0.97 (0.26-3.64)<br>0.9607 | 0.85 (0.21-3.50)<br>0.8268 | 1.35 (0.45-4.11)<br>0.5935 | 1.12 (0.35-3.55)<br>0.8482 |

**Table S3.** Long-term survival of germ cell tumors and thymomas, stratified by complexity score and primary vs. metastatic site.

| | | | N patients | 5-year Cumulative survival | Log-Rank P-value | 10-year cumulative survival | Log-Rank P-value |
|---|---|---|---|---|---|---|---|
| **Germ Cell** | | | | | | | |
| | | **Primary** | 65 | 75% | 0.5868 | 73% | 0.8168 |
| | | **Metastatic** | 140 | 76% | | 71% | |
| | **Complexity score L** | **Primary** | 11 | 89% | 0.4602 | 71% | 0.7901 |
| | | **Metastatic** | 132 | 79% | | 74% | |
| | **Complexity score I** | **Primary** | 6 | 67% | 0.0942 | 67% | 0.0942 |
| | | **Metastatic** | 3 | 0% | | 0% | |
| | **Complexity score H** | **Primary** | 48 | 72% | 0.0829 | 72% | 0.0829 |
| | | **Metastatic** | 5 | 40% | | 40% | |

| | | | N patients | 5-year Cumulative survival | Log-Rank P-value | 10-year cumulative survival | Log-Rank P-value |
|---|---|---|---|---|---|---|---|
| **Thymoma** | | | | | | | |
| | | **Primary** | 195 | 87% | 0.0501 | 65% | 0.0700 |
| | | **Metastatic** | 17 | 63% | | 52% | |
| | **Complexity score L** | **Primary** | 20 | 90% | 0.1652 | 72% | 0.2828 |
| | | **Metastatic** | 12 | 60% | | 60% | |
| | **Complexity score I** | **Primary** | 46 | 90% | 0.0744 | 66% | 0.0744 |
| | | **Metastatic** | 4 | 50% | | 50% | |
| | **Complexity score H** | **Primary** | 129 | 85% | 0.6880 | 64% | 0.0585 |
| | | **Metastatic** | 1 | 100% | | 0% | |

**Table S4**: Multivariate Cox regression models for 5-year and 10-year mortality: Model A adjusted for age and sex ; Model B adjusted for age, sex and type of tumor.

| | | **5-year survival**<br>*HR (95%CI)*<br>*p-value* | | **10-year survival**<br>*HR (95%CI)*<br>*p-value* | |
|---|---|---|---|---|---|
| | | **Model A** | **Model B** | **Model A** | **Model B** |
| **Complexity score** | | | | | |
| | **L** | Ref. | Ref. | Ref. | Ref. |
| | **I** | 0.94 (0.59-1.51)<br>0.8099 | 1.29 (0.79-2.11)<br>0.3104 | 0.97 (0.64-1.47)<br>0.8986 | 1.22 (0.80-1.88)<br>0.3587 |
| | **H** | 1.53 (0.98-2.37)<br>0.0604 | **2.66 (1.72-4.12)**<br>**<0.0001** | 1.36 (0.91-2.03)<br>0.1345 | **2.21 (1.48-3.30)**<br>**0.0001** |
| **Reconstructions** | | | | | |
| | **None** | Ref. | Ref. | Ref. | Ref. |
| | **1 OR 2** | 0.68 (0.44-1.04)<br>0.0753 | 0.71 (0.47-1.08)<br>0.1091 | 0.75 (0.51-1.11)<br>0.1559 | 0.72 (0.49-1.05)<br>0.0885 |
| | **≥ 3** | **1.66 (1.12-2.48)**<br>**0.0126** | 0.99 (0.66-1.47)<br>0.9419 | **1.75 (1.21-2.53)**<br>**0.0029** | 1.10 (0.76-1.60)<br>0.6233 |
